# Social vulnerability to health impacts of climate change in Australia: understanding drivers and inequalities

**DOI:** 10.1101/2025.07.03.25330731

**Authors:** Ang Li, Mathew Toll, Erika Martino, Lisa Gibbs, Emma McNicol, Kate Mason, Rebecca Bentley

## Abstract

**Background:** A limited ability to identify social vulnerability and community resilience at local scales has been recognised as a critical barrier to both climate adaptation and health risk assessment and planning. This study aims to assess multi-domain social vulnerability to the health impacts of climate change across communities in Australia using a system based approach, quantify contributions across intervention levers, and identify key drivers of health inequalities.

**Methods:** Informed by a scoping review and the WHO Social Determinants of Health Equity framework, we compiled area-level data from multiple sources on 57 social vulnerability indicators, subsumed under 32 subdomains and 7 domains (demographic profile, economic security, residential environment, infrastructure and services, community context, population health, and governance and policies). These indicators were used to construct a Social Vulnerability Index for the Health Impact of Climate Change (SVI-HICC) and scores in each domain. We examined the association between the SVI-HICC and health outcomes related to extreme weather and climate events, evaluated its performance relative to alternative social indices, and identified main contributors to social vulnerability using dominance analysis.

**Findings:** Spatial mapping revealed geographic variation in the composition of social vulnerabilities across domains and subdomains. High social vulnerability was more common in regional and remote areas, primarily associated with demographic and economic disadvantages, while pockets of high social vulnerability were also observed in urban areas, largely attributable to residential environment and community context. People in highly vulnerable areas experienced greater losses in mental, emotional, social, physical, and general health from weather and climate extreme events. Community context (driven by social support and networks), residential environment (driven by dwelling condition), and infrastructure and services (driven by communication access) were identified as the strongest contributors to social vulnerability.

**Conclusion:** An area-level assessment of social vulnerability makes visible how social and structural determinants contribute to health inequities in climate change. Such insights can inform climate adaptation policies that are equity-oriented and context-sensitive.

## Introduction

The impacts of climate change are increasingly documented across human health and broader social systems ^1^. Anthropogenic climate change is exerting both direct and indirect effects on health, that are projected to worsen without serious mitigation and adaptation efforts ^1–3^. The scale of harm to human health has led *The Lancet Countdown* to conclude that climate change is affecting all monitored domains of human health ^2^. These health burdens are, however, not evenly distributed across population groups. Health risks from climate change emerge from the intersection between climate hazards, exposure, and vulnerabilities ^1^. Although climate change exposes all populations to environmental hazards, the degree of susceptibility to its health impacts varies considerably across communities, as social determinants can significantly mediate or moderate health risks ^4^.

The recent *Intergovernmental Panel on Climate Change* (IPCC) report underscored the importance of understanding social vulnerability at the community level, noting that a “limited ability to identify social vulnerability and community strengths” at local and urban scales constitutes a critical barrier to effective climate adaptation ^1^. Mapping the geographic distribution of hazards and exposure is a necessary but insufficient step in assessing climate-related health risk. Mitigating health risks from climate change requires an assessment of local social vulnerability to identify populations at heightened risks, to understand the underlying drivers of vulnerability, and to inform equitable adaptation planning aimed at strengthening community resilience.

Social vulnerability to the health impacts of climate change is shaped by a wide range of individual, structural, and institutional factors that generate relative group or community advantage and disadvantage. An expanding corpus of research has examined determinants such as demographic structures (e.g., age, sex, and ethnicity), socioeconomic status (e.g., income, employment, and education), and, to a lesser extent, access to infrastructure and services ^5,8^. Many of these factors are commonly integrated into social vulnerability indices (SVIs), one of the main tools to assess susceptibility on a local scale, especially in disaster management, to identify susceptibility to environmental hazards ^6,7,9^.

However, as indicated in several reviews and critiques of the existing literature on social vulnerability indicators and indexes ^5,10^, the operationalisation of social vulnerability has frequently depended on a narrow set of demographic and socioeconomic attributes, which do not fully capture the conditions in which group membership translates into susceptibility ^5^. Overreliance on variables that capture individual demographic characteristics is akin to analysing pre-constructed ideas and not delving into the drivers and systematic causes of vulnerability ^5,10^. Categories of sex and remoteness, for instance, are only indicative of incapacity under certain social and institutional arrangements that produce disadvantage for a particular gender or remoteness category. Broader social determinants – such as governance, housing adequacy, and social inclusion – that reflect social, political, and institutional contexts are necessary to explain the conditions under which vulnerability emerges but are often overlooked in assessments ^5,10–12^ .

In addition, despite growing recognition of climate change as one of the greatest threats to human health, the development and application of social vulnerability metrics specifically designed to assess health risks of climate change remain limited ^5,8^. While SVIs have been applied by policymakers to direct resources towards vulnerable groups in response to public health crises and disasters ^13–15^, few index-based studies have been explicitly linked to health measures, or examined domain specific drivers contribute to vulnerability to climate health risks ^8^. Many recent studies in the fields of climate change, disaster management, and health replicate existing social vulnerability indices developed for other domains without adapting it to or validating it in the context of climate change and health ^8,16^. Reviews of social and health vulnerability assessments found that studies often lacked justification for their adopted methodology or validation of their approach to identifying vulnerabilities ^10,17^. There is a need for SVIs that are designed for understanding climate-related health risks.

These methodological gaps are further compounded by the limited engagement with structurally grounded theories in much of the existing research on social vulnerability and resilience indices ^18^. Studies that incorporate theory tend to focus narrowly on explanations of individual behaviour or risk perception, overlooking broader institutional and structural dimensions of vulnerability ^11,18^. This relative neglect of relational framing of social vulnerability, alongside a lack of data infrastructure, results in the paucity of structural and institutional indicators in SVIs ^5^. Inclusion of institutional and structural indicators, alongside contributing to an understanding of how group vulnerability is produced, provide a means to move beyond simple composite SVI scores and decompose vulnerability into multiple domain contributions that can be used to identify leverageable intervention points.

This study aims to develop an area-based multi-domain system-based Social Vulnerability Index for the Health Impact of Climate Change (SVI-HICC) in Australia – a country amongst the most exposed to climate change. This study outlines the conceptualisation of social vulnerability to climate health risks, describes and validates the construction of the SVI-HICC comprised of multiple domains and subdomains, and identifies key drivers of health risks from climate change and their relative contributions. The conception of social vulnerability, selection of indicators, and validation against health outcomes from climate-related exposures seek to overcome many of the critiques associated with prior SVI developments ^5,10^. Through this, we illustrate how SVI-HICC provides a tool to assess how social factors across domains contribute to susceptibility to climate risks and pinpoint leverage points to reduce that susceptibility.

## Methodology

### Conceptual Framework

Social vulnerability is rooted in social and place-based inequality ^6,7^. To conceptualise the link between social vulnerability and unequal health impacts from climate change, this study adapts the WHO’s Social Determinants of Health Equity (SDHE) framework ^19^ and proposes measurements guided by a scoping review of social vulnerability indicators ^5^. Informed by the conceptual framework and literature synthesis of indicators, social vulnerability was disaggregated into multiple domains and subdomains (Figure 1). These encompass factors that reflect broader structural determinants of health, diverse socioeconomic systems, and various forms of disadvantage and marginalisation that harm health, and can be operationalised to assess social determinants of health inequities ^19^. The SVI-HICC is constructed from indicators (57) grouped into subdomains (32) nested within principal domains (7). Together, these domains and subdomains form a multi-domain, system based index, where each domain and subdomain represents a distinct but interrelated facet of social vulnerability.

**Figure 1.**
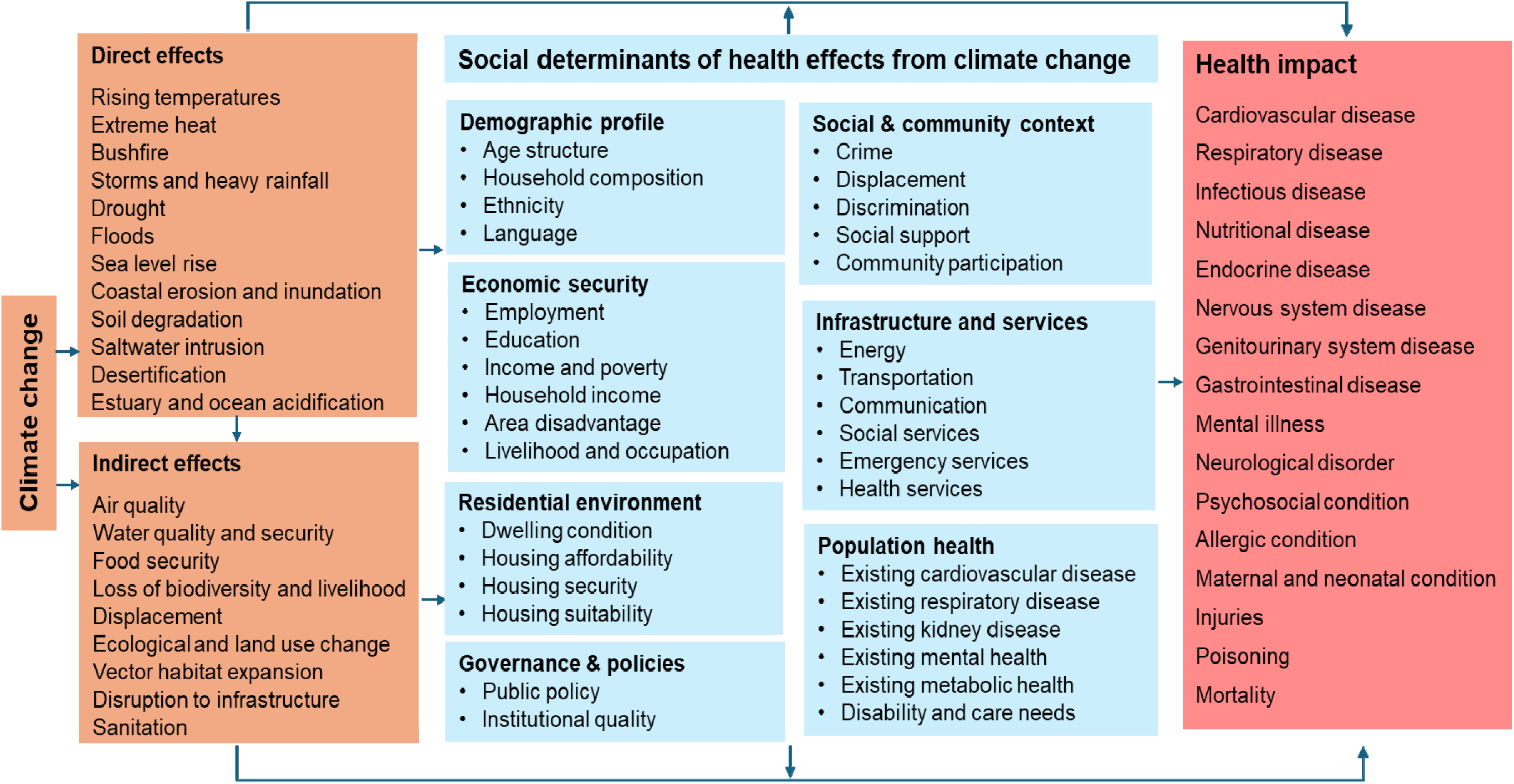
Conceptual Framework on social vulnerability to the health impacts of climate change Notes: The direct and indirect links between climate change hazards and health impacts are drawn from previous reviews and reports ^1,3,5^. Some of the well-established health impacts include cardiovascular conditions linked to storms, hurricanes and cyclones (e.g., Hurricane Katrina in 2005 and Hurricane Sandy in 2012 off the coast of the US), respiratory diseases from wildfires (e.g., the Australian Black Summer bushfires in 2019 and Northern California wildfires in 2017), and heat-related morbidity and mortality associated with heatwaves (e.g., more frequent and severe extreme heat impacting countries globally), among others.

### Data

Data for the SVI-HICC was compiled from multiple publicly available data sources (see Table 1). The unit of data was at the Statistical Area Level 2 (SA2), designed to represent communities that interact socially and economically together by the Australian Bureau of Statistics. There are 2,454 spatial SA2s in Australia, with an average population of 10,000. Data at other geographic levels (e.g., local government areas) were transformed to the SA2 level for consistency using the ABS correspondence files. When inferring population estimates from nationally representative samples, population weights were applied ^21^. The SVI-HICC was constructed for census years 2016 and 2021.

**Table 1.**
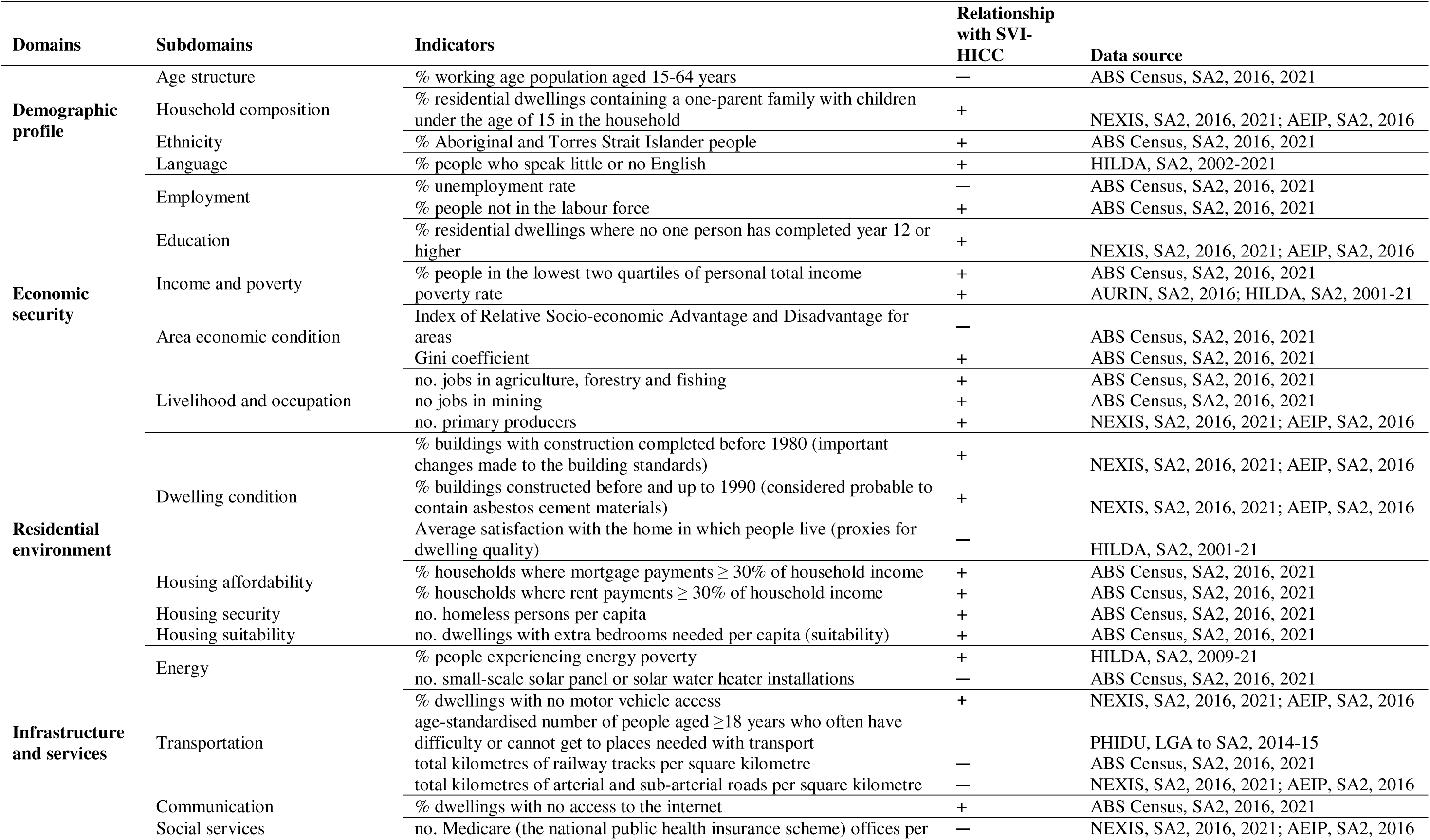
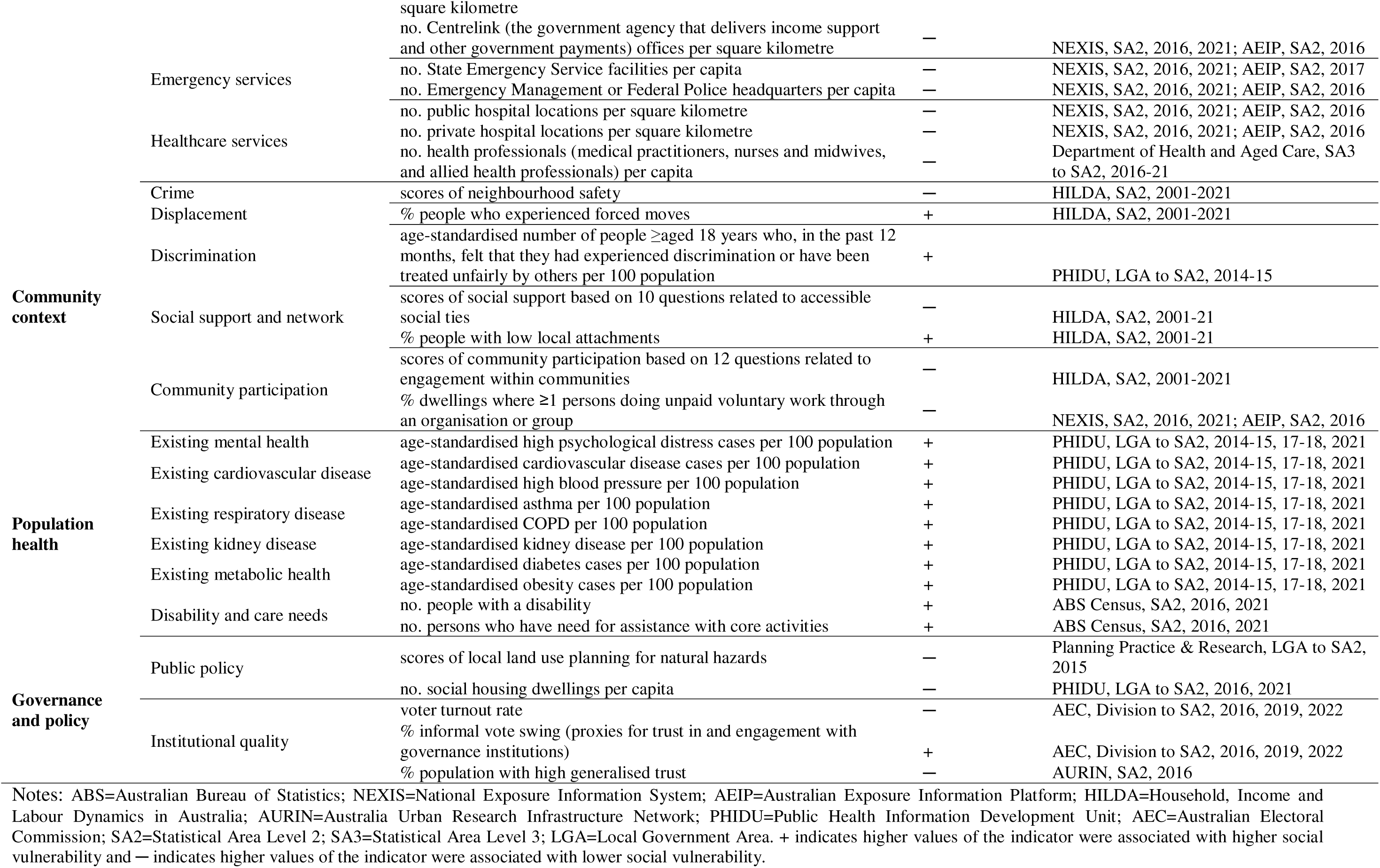
Domains, subdomains, and indicators included in the SVI-HICC.

### Domain and subdomain identification

Drawing on evidence from the literature review and adapting the SDHE framework, domains and subdomains were identified for inclusion based on their relevance to the Australian context, regional variability, and availability of data. Seven conceptually distinct domains were developed, including demographic profile, economic security, residential environment, infrastructure and services, community context, population health, and governance and policy. Each domain consists of subdomains with established impacts on health equity ^19^.

### Indicator selection

A comprehensive search was conducted to identify datasets containing relevant indicators. The selection of indicators was based on their conceptual alignment with the literature review, theoretical framework, and scope for empirical operationalisation ^5,23^. These processes buttressed the theoretical coherence, internal consistency and robustness, practical applicability, and conceptual relevance of the indicators ^23^.

### Index construction

In line with the framework developed in Figure 1, the SVI-HICC was constructed as a top-down hierarchical model, with indicators grouped into subdomains and domains and standarised using percentile ranking ^9,24^. Alternative approach was employed to test the sensitivity of the index construction using a principal component analysis (PCA) method (see Appendix for details). Data processing and analyses were conducted using Stata 18.0, and mapping was generated using Datawrapper.

### Validity assessment

The validity and reliability of the SVI-HICC was assessed following the OECD framework for composite indicators ^20^ and the psychometric assessment framework for constructs ^26^ that consider translational validity and criterion validity ^14^ (see Appendix for details). These assessments were conducted by 1) assessing the framework and literature review guiding the construction of indicators, subdomains, domains, and the index, 2) testing the Pearson correlation and Spearman correlation between SVI-HICC and health outcomes from climate extreme events (measured using mental, social, emotional, and physical health domains of the 36-Item Short Form Survey (SF-36) when exposed to floods, bushfires, and cyclones, available in the HILDA Survey), and 3) comparing the performance of the SVI-HICC with generic socio-economic indexes for areas developed by the ABS including the Index of relative socio-economic advantage and disadvantage (IRSAD) and Index of economic resources (IER) standardised by min-max scaling.

### Relative contribution of subdomains and domains to health risks from climate change

Dominance analysis ^27^ was conducted to determine the relative importance of subdomains (within each domain) and domains (within the overall index) in explaining community health vulnerabilities to climate change. Specifically, dominance analysis quantifies the contribution of each vulnerability component by assessing the reduction in prediction error attributable to that component in statistical models where the dependent variables were climate-related health risks, as defined above.

## Results

Figure 2 maps the distribution of the overall and domain-specific indices. The composition of social vulnerabilities in each domain differs across areas. While some areas experienced consistently high vulnerability across multiple domains, others displayed more domain specific vulnerabilities. Higher social vulnerability is more common in regional and rural areas (the mean of SVI-HICC deciles=7.03) than in capital cities (the mean of SVI-HICC deciles=4.55), particularly in the demographic and economic domains, though rural areas have lower social vulnerabilities in the domains of residential environment and community context. Some domains are more clustered (e.g., demographic domains in the Northern Territory) than others. The distribution of subdomains also differs across areas. For instance, some areas exhibited greater physical health vulnerability while others experienced greater mental health vulnerability, and housing unaffordability did not necessarily overlap with housing insecurity (homelessness), illustrating the diverse pathways of vulnerability across communities.

**Figure 2.**
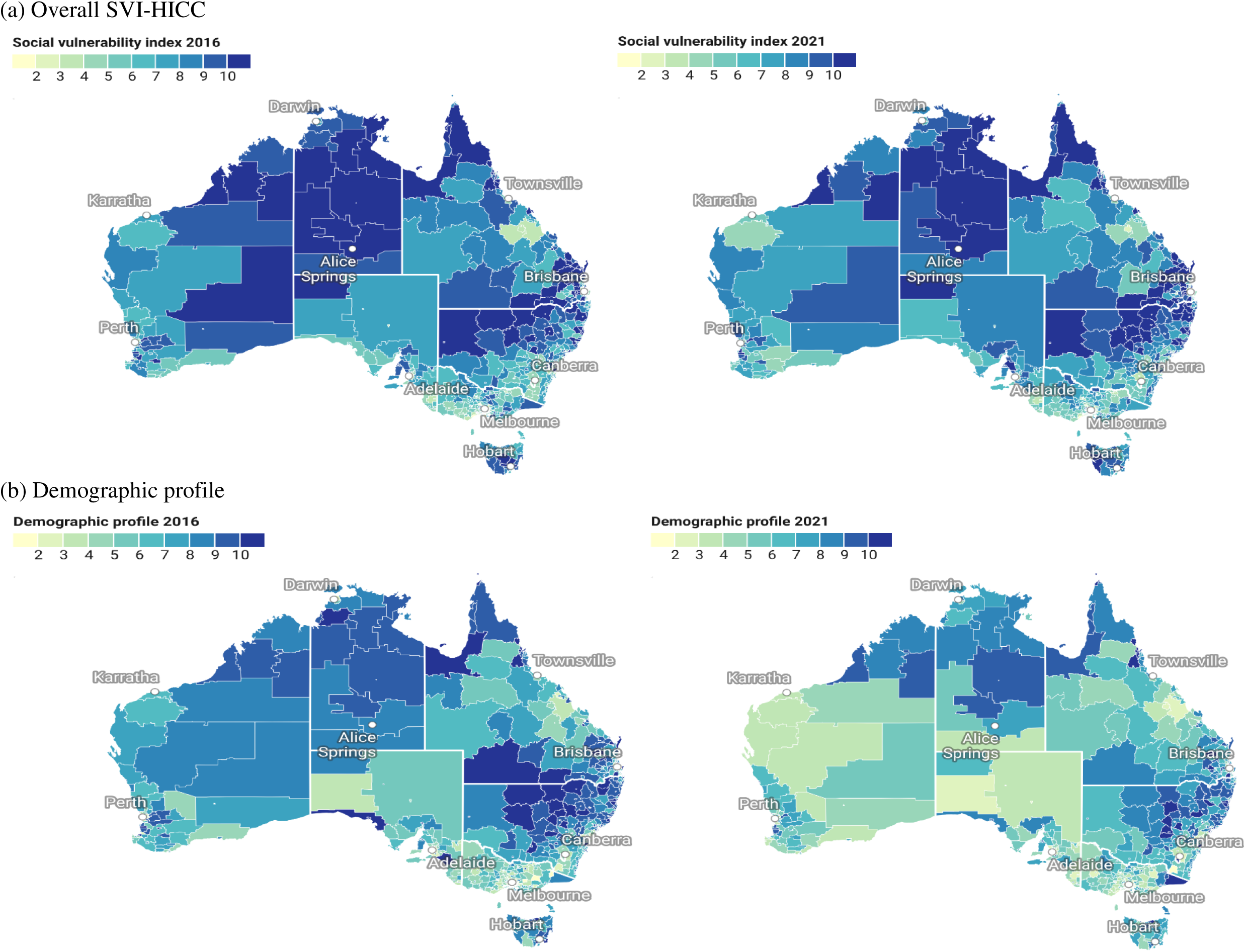
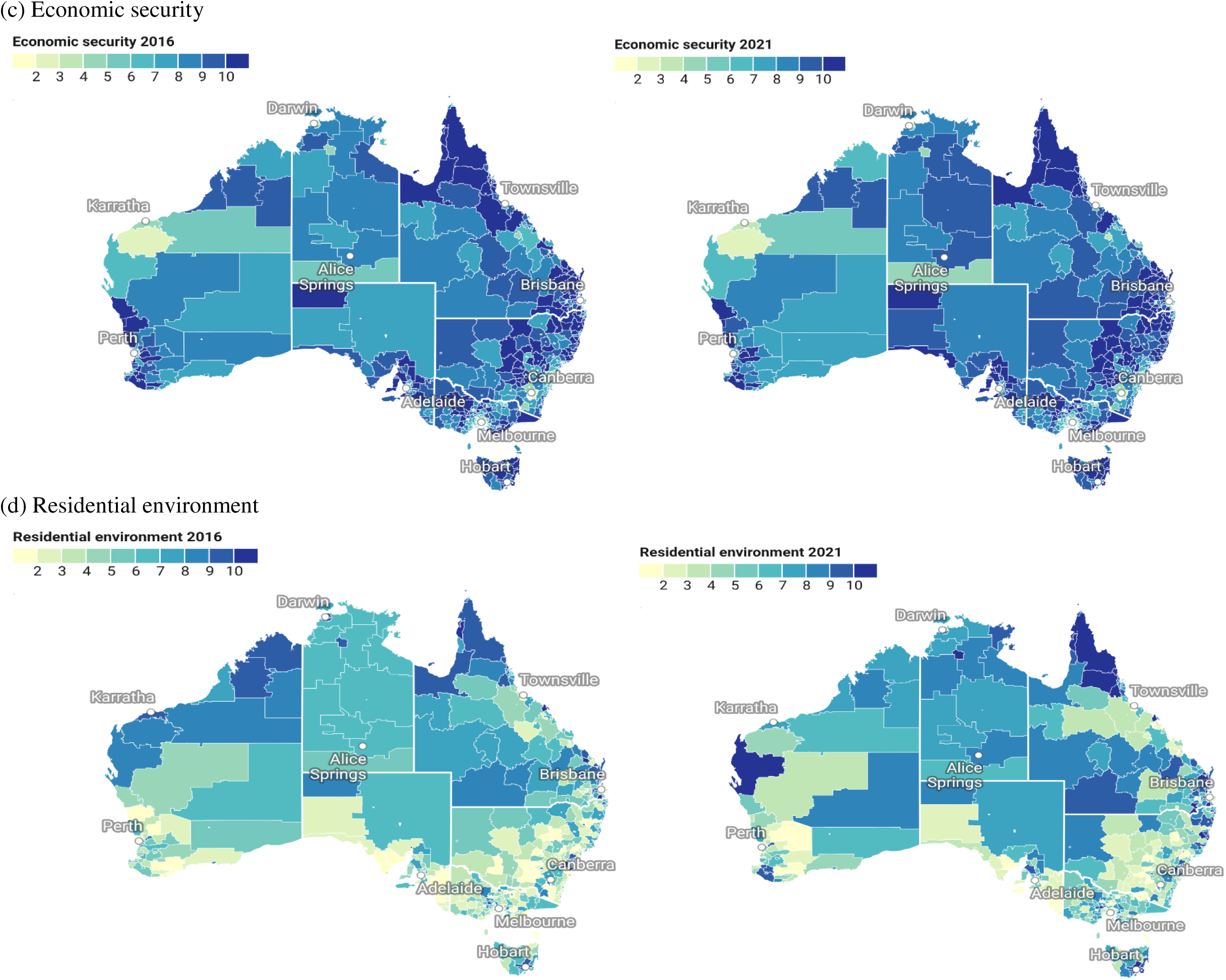
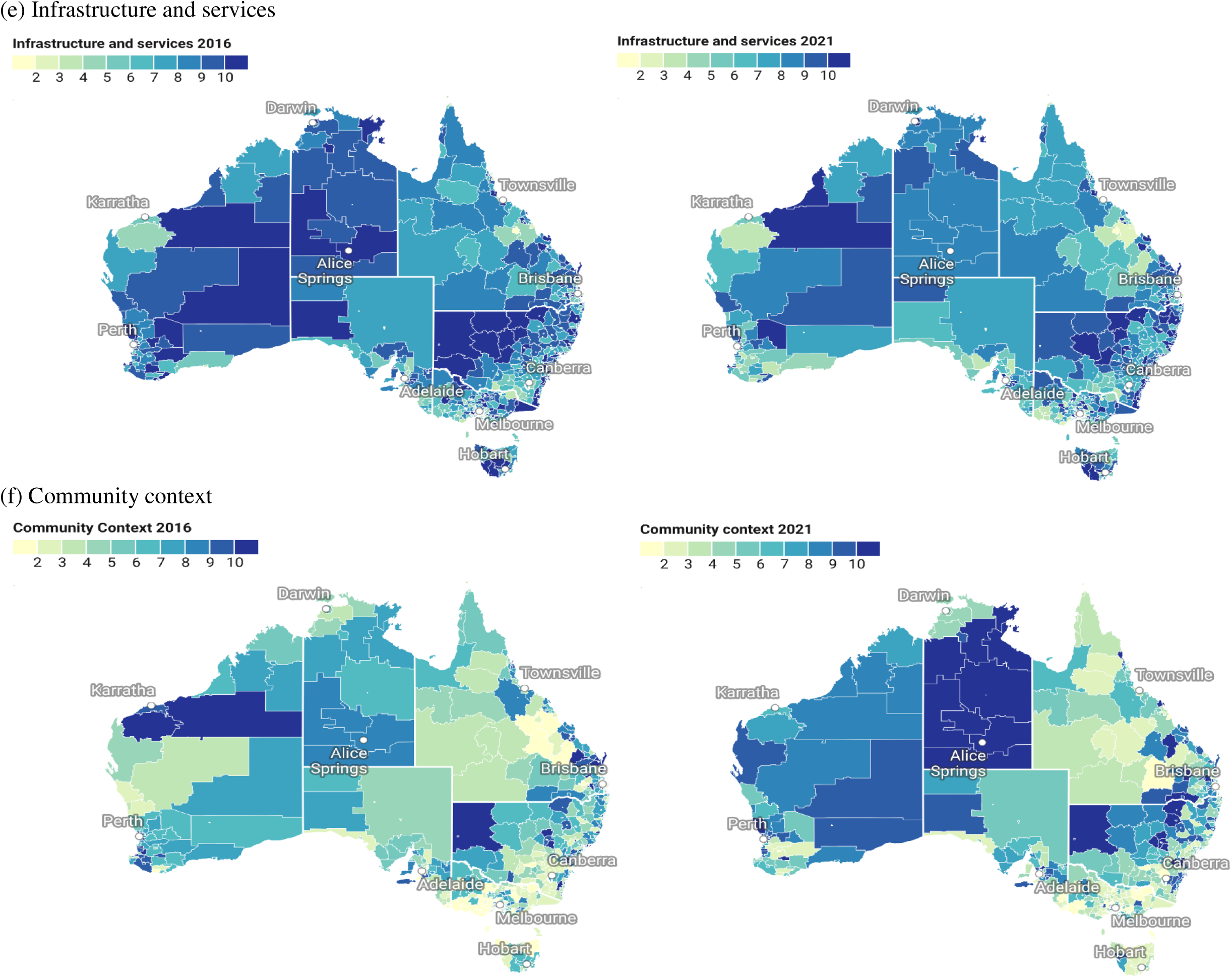
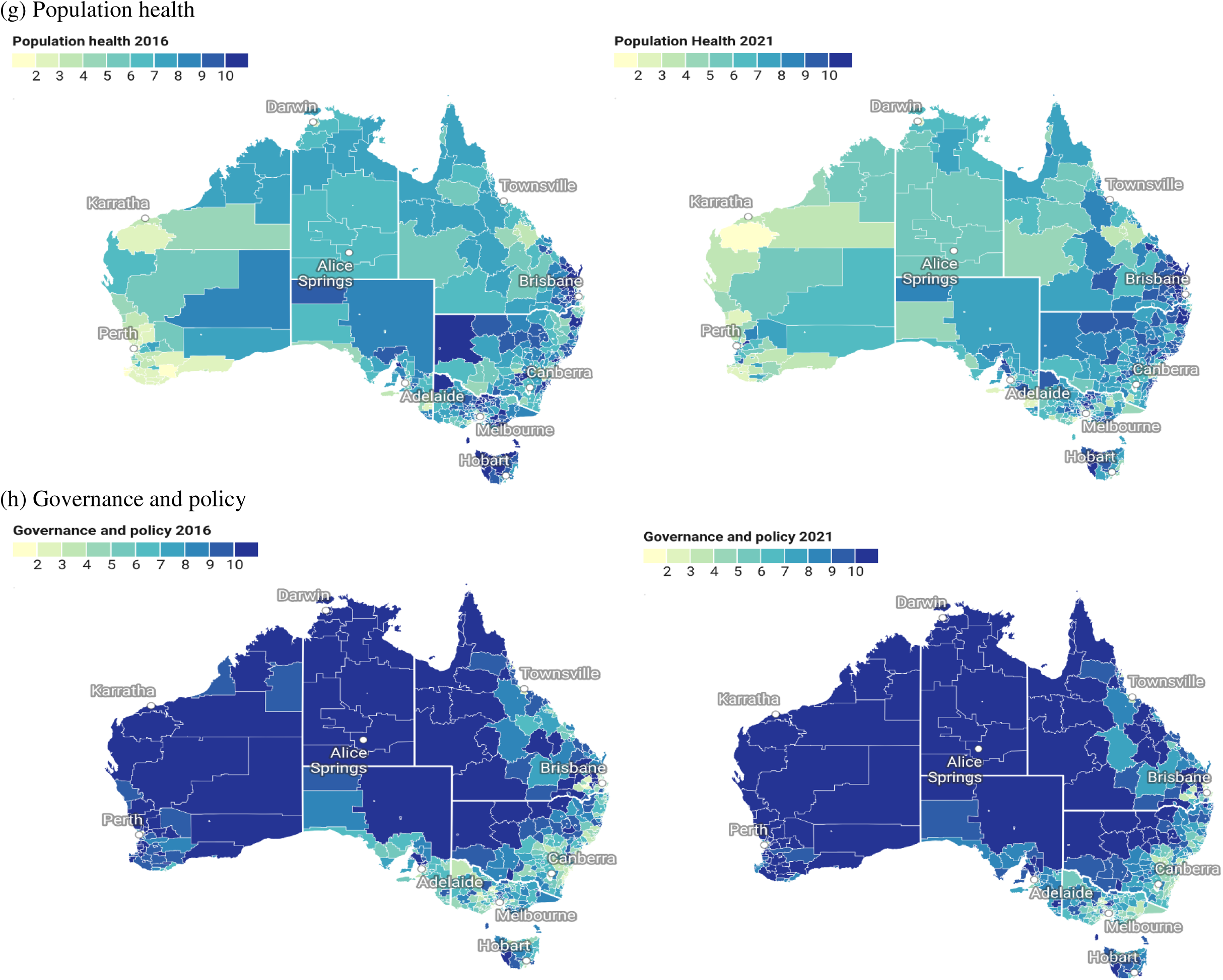
Geographic mapping of the SVI-HICC

In assessing validity and reliability, the alignment of the SVI-HICC with the WHO’s SDHE Framework ^19^, supported by an extensive scoping review ^5^ and informed by discussions and consultations with subject matter experts, provides a basis for establishing translational validity. To verify criterion validity, the SVI-HICC demonstrated strong associations with climate related health outcomes, with Pearson correlation and Spearman correlation markedly greater than those observed for generic socioeconomic indices (including IRSAD and IER), indicating its greater relevance for capturing climate-related health vulnerability (Table 2). The alternative index derived using PCA was highly correlated with the SVI-HICC and showed consistent associations with climate-related health outcomes.

**Table 2.** Associations between the SVI-HICC and climate-related health outcomes.

|  | Mental<br>health | Emotional<br>wellbeing | Social<br>functioning | Physical<br>functioning | General<br>health |
| --- | --- | --- | --- | --- | --- |
| **Pearson correlation** |  |  |  |  |  |
| SVI-HICC | -0.214*** | -0.205*** | -0.203*** | -0.176*** | -0.204*** |
| Index of Economic Resources | -0.108* | -0.119* | -0.094 | -0.122** | -0.076 |
| Index of Relative Socio-economic Advantage/Disadvantage | -0.145** | -0.131** | -0.141** | -0.191*** | -0.136** |
| **Spearman correlation** |  |  |  |  |  |
| SVI-HICC | -0.207*** | -0.184*** | -0.197*** | -0.181*** | -0.193*** |
| Index of Economic Resources | -0.137 | -0.120 | -0.092 | -0.125 | -0.083 |
| Index of Relative Socio-economic Advantage/Disadvantage | -0.162*** | -0.133** | -0.141** | -0.176*** | -0.138** |

Notes: All indices were standardised using min-max scaling and oriented in the same direction, with higher scores indicating higher vulnerabilities or disadvantages. \*\*\*p<0.01, \*\*p<0.05, \*p<0.10.

Results from linear regression models showed that each one-point increase in the SVI-HICC (scaled from 0 to 100) was associated with decreases of 0.2, 0.3, 0.4, 0.2, and 0.2 points on average in mental health, social functioning, emotional wellbeing, physical functioning, and general health among affected populations following the extreme events, respectively (Figure 3). These estimates imply that communities differing by 10 points (one decile) on the SVI-HICC would differ by 2-4 points in health-related quality of life outcomes, while a 20-point difference (one quintile) would correspond to differences of 4-8 points.

**Figure 3.**
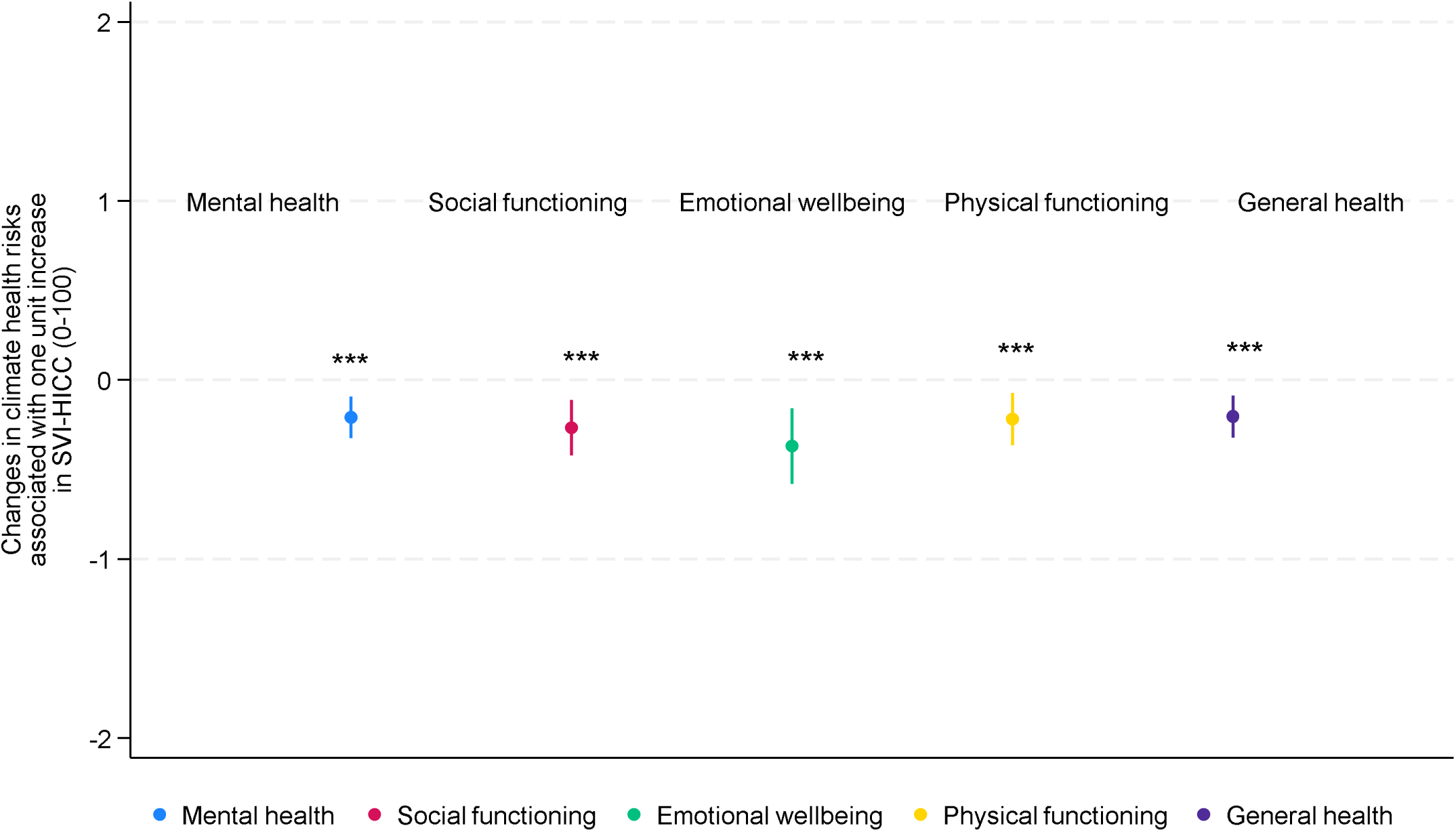
Association between SVI-HICC and climate-related health outcomes Notes: Health outcomes associated with extreme weather and climate events were regressed on the SVI-HICC (scaled from 0 to 100) using linear regression models. ***p<0.01.

Figure 4 illustrates the relative importance of each subdomain and domain within the SVI-HICC in contributing to the health effects from extreme climate events using dominance analyses. Across domains, community context explained the most variance in climate-related health outcomes, followed (in order of relative importance) by residential environment, infrastructure and services, population health, demographic profile, economic security, and governance and policy. For each local area, high scores in lower-ranking domains (e.g., demographic profile) could be offset by low scores in higher-ranking domains (e.g., residential environment) contributing to climate risks to health. Within each domain, vulnerability levels were influenced by the differential contributions from constituent subdomains. The community context domain was most influenced by the extent of social support and network. For the residential environment domain, dwelling conditions were central in shaping health-related inequalities. In the infrastructure and services domain, communication was the major contributor. With the population health domain, disability and care needs had the strongest impact. For demographic profile, economic security, and governance domains, multiple subdomains were similarly influential.

**Figure 4.**
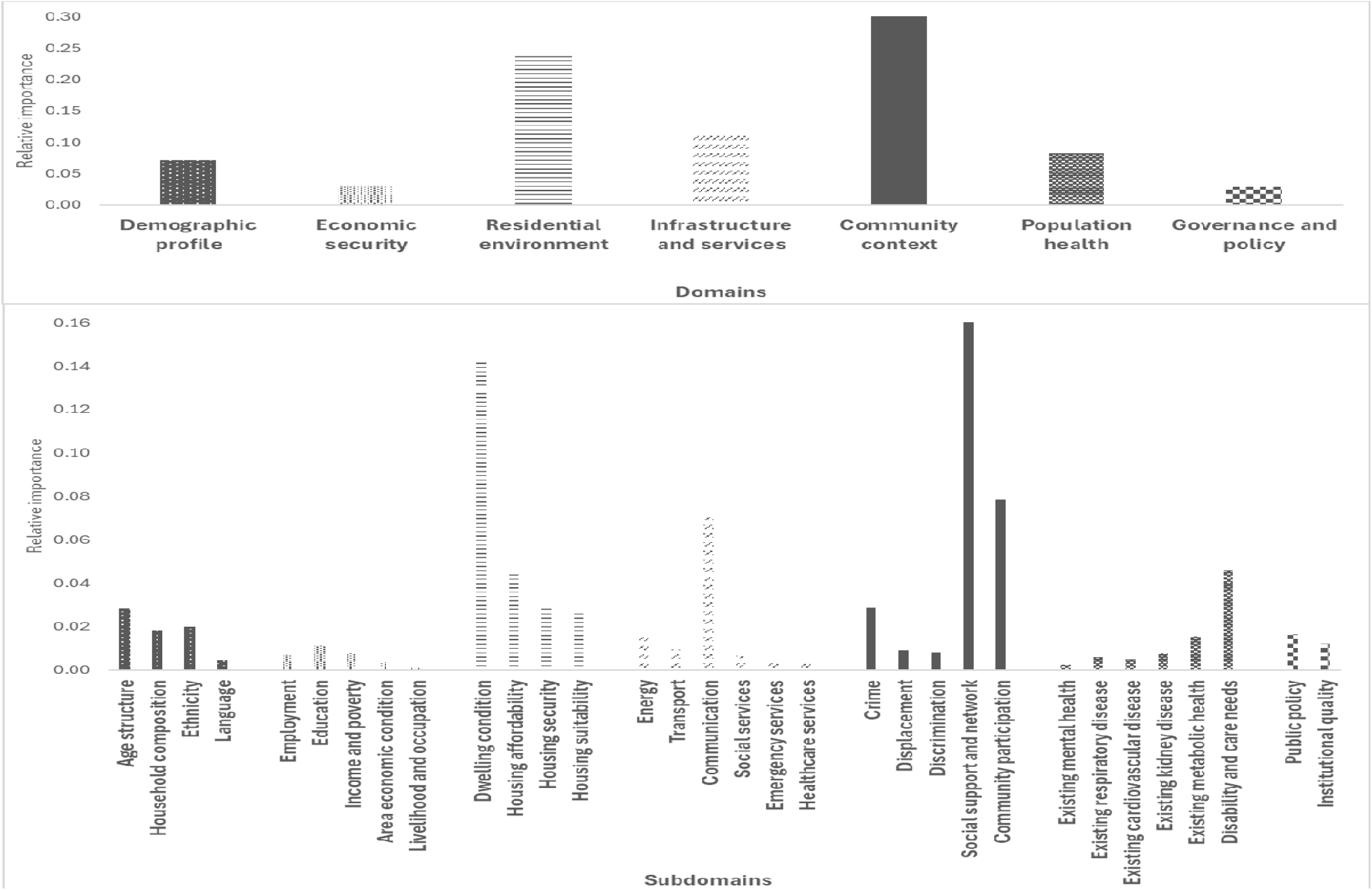
Relative contribution of social vulnerability subdomains and domains to the health effects from climate change Notes: The relative importance of domains within the SVI-HICC (domain weights, which are summed to 1 across all domains) is presented on the top panel, and the relative importance of subdomains within each domain (subdomain weights within each domain multiplied by the corresponding domain weight, which are summed to 1 across all subdomains) is presented on the bottom panel.

## Discussion

This study presents an assessment of social vulnerability to the health impacts of climate change across communities in Australia through the development and operationalisation of a multi-domain, system-based index (SVI-HICC) that captures diverse dimensions of and contributions to vulnerability. By integrating a conceptual framework with evidence from a scoping review to inform the selection of indicators, subdomains, and domains, this study demonstrates disparities in health impacts from climate change across varying levels of social vulnerability and identifies the main drivers of vulnerability across domains and subdomains. The design and construction of social vulnerability measures within a holistic and integrated framework demonstrate that the SVI-HICC – explicitly and conceptually grounded in the SDHE framework within the context of climate change – presents greater relevance in assessing social vulnerabilities to health effects of climate-related events than generic socioeconomic indices. In doing so, this study offers a spatially and structurally grounded tool for understanding and addressing inequities in climate health risks.

The SVI-HICC revealed substantial geographic variation in relative climate health vulnerability across Australia. High vulnerability is more concentrated in regional and remote areas, driven by demographic profile and economic security, with pockets of high vulnerability also present within metropolitan areas, driven by residential environment and community context. Social vulnerability is multidimensional and context specific. The distribution of domains and subdomains differs across areas, and the composition of social vulnerabilities varies across communities, suggesting that communities experienced climate-related health vulnerabilities through different combinations of underlying determinants depending on their locations. Geographic variation in domain-specific vulnerabilities highlights the importance of place-based adaptation strategies.

The extent of health risks from climate change experienced by individuals was significantly associated with the level of SVI-HICC, with individuals in areas of higher social vulnerability experiencing substantially poorer mental, emotional, physical, social, and general health following extreme weather and climate events. These unequal health patterns underscore the critical role of social vulnerability in shaping the distribution of climate-related health burdens, and, importantly, the need to identify and support at-risk communities through a systems-based approach that addresses vulnerability across diverse social spheres and geographic regions.

Findings also point to the need to address structural inequities to prevent the exacerbation of existing health disparities. The more influential domains in explaining social vulnerability to short-term impact of extreme weather and climate events were community context, residential environment, infrastructure and services, and population health, and, to a lesser extent, demographic profile, economic security, and governance and policy. This highlights the importance of delving more into the structural dimensions of social vulnerability in empirical studies and health risk assessment ^5,12^, but also provides a silver lining, as the more salient SVI-HICC domains can be ameliorated with policy action. This further underscores that some policy levers which address the health impacts of climate change sit outside of the health portfolio and the need for a Health in All Policies approach to effective and equitable climate adaptation.

Major subdomain contributors underpinning these domain-level vulnerabilities include social support and network, housing condition, communication, community participation, and disability and care needs. This aligns with emerging evidence that demonstrates the role of these subdomains in shaping climate-related health risks for individuals and communities. Individuals experiencing housing insecurity and those with disabilities were more vulnerable to adverse health impacts following climate-related disasters ^28–30^, while strong social support and community participation were associated with greater health resilience ^31,32^.

This study makes several important contributions. Existing studies on social vulnerability to the health impact of climate change often depend on a narrow operationalisation of vulnerability, frequently overlooking the effect of broader social determinants of health across domains ^5^. The SVI-HICC addresses this limitation by incorporating a broader range of social indicators, enabling a more comprehensive and valid assessment of social vulnerability. With few existing SVIs specifically focused on the intersection of climate change and health ^8^, this study contributes to methodological and empirical development in this area. By linking to health impacts from climate change, this research provides a tool to understand how social determinants across multiple domains and subdomains shaped health risks from climate change and to identify the individual and structural drivers of climate-related health inequities.

The study generated insights to help identify leverage points for interventions and support planning, adaptation, and resilience building efforts. The SVI-HICC can be used to introduce and incorporate new datasets and to measure changes over time. In conjunction with information on hazard exposures to triangulate areas at risk of climate-related health impacts, this tool can be used to assess where, how, and the extent to which climate change poses health risks and identify the social domains that contribute to susceptibility. The multi-domain approach applied in this study provides a foundation for evidence-based, equitable policy design for climate adaptation planning and social vulnerability reduction. The SVI-HICC can be leveraged to guide placed-based planning and allocation of funding, resources, and services for governments across levels, facilitate council planners in developing locally relevant interventions (e.g., nature based solutions and urban renewal projects), and strengthen collaboration among community organisations forming alliances and building local capacity by enabling identification of common issues and interests. By adopting a Health in All Policies perspective, the tool supports community health and resilience through proactive prevention and mitigation of climate-related risks, informed by local vulnerability drivers and exposure contexts.

Further research could extend this study in several ways. First, improving the health specificity of the index by linking social vulnerability more directly to specific health outcomes, such as cardiovascular disease, kidney disease, or mental health, would enable more targeted and effective health responses. Second, the availability and granularity of indicators could be expanded to better capture structural dimensions of social vulnerability to climate risks such as organisational governance, climate preparedness and literacy, emergency education and supplies, and dwelling resilience features ^5,15^. Third, current validation and health inequality analyses focused on rapid-onset events and short-term impacts. Further research could extend this work to slow-onset events such as droughts and projected impacts under future climate change scenarios.

## Conclusion

Understanding social vulnerability is a critical component of effective planning for and mitigating the health effects of climate change. This study assessed multi-dimensional social vulnerability, with subdomains and domains, across local communities in Australia, and identified key drivers of climate health risks and inequities. Findings show that individuals in areas of higher social vulnerability experienced greater health loss associated with exposure to climate-related extreme events. The variation in vulnerability and its underlying drivers across geographic areas and time necessitates planning and policy responses that are equity-oriented and context-sensitive. This multi-domain assessment of social vulnerability can be paired with area-level mapping of climate hazards and exposures to help inform climate health risk assessments and adaptation planning to mitigate the health losses associated with climate change.

## Data Availability

All data produced are available online at https://dbr.abs.gov.au/ and https://doi.org/10.26193/24EJST.

## Data sharing statement

Data supporting this study are publicly available from the Australian Bureau of Statistics, ADA Dataverse, Geoscience Australia, Australia Urban Research Infrastructure Network, Public Health Information Development Unit, Australian Electoral Commission, and Department of Health and Aged Care.

## Acknowledgments

The research was conducted with funding support from the Commonwealth Climate Resilience Network Grant and the Australian Research Council Discovery Early Career Researcher Award (Grant number DE240101135). An early version of this work was presented at the Annual Conference of the International Society for Environmental Epidemiology (Chile, 2024). We would like to thank colleagues from the community and government organisations for their valuable discussions and Dr Annabelle Workman from the University of Melbourne for their insights and feedback.

